# A Real-World Retrospective Study of Sintilimab in Combination with Neoadjuvant Chemotherapy for Triple-Negative Breast Cancer

**DOI:** 10.64898/2026.04.02.26349910

**Authors:** ZhaoBing Gao, HaiWen Liang, Xin Bai, Kang Dong, Jing Li, WanJia Qiao, BiaoFeng Shan, XiaoHua Chen, JianMing Tang

**Affiliations:** The First Clinical Medical College, Lanzhou University, Lanzhou, Gansu, China; Department of Radiation Oncology, The First Hospital of Lanzhou University, Lanzhou University, Lanzhou, Gansu, China

**Keywords:** Triple Negative Breast Cancer, Immunotherapy, Sintilimab, Combination neoadjuvant chemotherapy, Efficacy, Real-World data

## Abstract

**Purpose:** This study aimed to evaluate the efficacy and safety of neoadjuvant chemotherapy (NAC) combined with the programmed death protein 1 (PD-1) inhibitor sintilimab versus NAC alone in patients with triple-negative breast cancer (TNBC).

**Materials and Methods:** In this retrospective cohort study, we collected clinical data from 61 patients with triple-negative breast cancer (TNBC) who received neoadjuvant therapy at The First Hospital of Lanzhou University between July 2024 and July 2025. These patients were divided into two groups: the neoadjuvant chemotherapy (NAC) plus sintilimab group (n=27) and the NAC-alone group (n=34). The primary endpoint was the pathological complete response (pCR) rate. Secondary endpoints included objective response rate (ORR), safety, and changes in tumor markers.

**Results:** The combination therapy group showed significantly higher ORR (85.2% vs. 58.8%) and pCR rates (59.3% vs. 32.4%) compared to the NAC alone group (both P<0.05). Post-treatment Ki-67 levels were also significantly lower in the combination group (P<0.05). The overall incidence of adverse events was comparable between groups (P>0.05), although leukopenia was more frequent with sintilimab (P<0.05).

**Conclusion:** In the neoadjuvant setting for TNBC, the addition of sintilimab to NAC significantly improves ORR and pCR rates, effectively reduces the tumor proliferation index Ki-67, and does not significantly increase the overall burden of adverse events. The combination regimen shows a manageable safety profile and demonstrates positive clinical value.

## Introduction

Triple-negative breast cancer (TNBC) is a breast cancer subtype characterized by the absence of estrogen receptor (ER), progesterone receptor (PR), and human epidermal growth factor receptor 2 (HER2) expression. It represents a major clinical challenge due to its aggressive behavior, high risk of recurrence, and the lack of well-defined molecular targets for therapy. [1] Anthracycline combined with taxane represents the standard neoadjuvant chemotherapy regimen for triple-negative breast cancer; however, the pathological complete response (pCR) rate appears to have plateaued, highlighting the urgent need for novel therapeutic strategies.[2]

In recent years, breakthroughs in cancer immunotherapy have transformed the treatment paradigm for triple-negative breast cancer (TNBC). Basic research has demonstrated that TNBC typically exhibits a higher tumor mutational burden and increased levels of tumor-infiltrating lymphocytes, providing a potential biological rationale for the application of immunotherapy.[3] Inhibitors targeting programmed death-1 and its ligand (PD-1/PD-L1) have demonstrated remarkable efficacy across various solid tumors by blocking the immunosuppressive signaling pathway and consequently reactivating T cell-mediated antitumor immunity.[4] The pivotal phase III KEYNOTE-522 trial demonstrated for the first time that adding pembrolizumab to standard neoadjuvant chemotherapy significantly improved both the pathological complete response rate and event-free survival in patients with triple-negative breast cancer, thereby establishing the combination of immunotherapy and chemotherapy as a cornerstone of neoadjuvant treatment.[5] However, the high cost and limited accessibility of imported agents such as pembrolizumab remain a significant barrier. This highlights the important clinical and practical value of exploring domestic PD-1 inhibitors, which may offer comparable efficacy at a more favorable cost-effectiveness profile.

Sintilimab is a fully human IgG4 monoclonal antibody that binds to PD-1, thereby blocking its interaction with PD-L1 and PD-L2. This mechanism helps restore endogenous antitumor T-cell responses. The drug has been approved by the China National Medical Products Administration for the treatment of classical Hodgkin lymphoma, non-small cell lung cancer, hepatocellular carcinoma, esophageal squamous cell carcinoma, and gastric or gastroesophageal junction adenocarcinoma.[6][7][8][9][10] In the field of breast cancer, particularly in the neoadjuvant treatment of triple-negative breast cancer, evidence for sintilimab in combination with chemotherapy is accumulating. Preliminary data from small-scale phase II studies have shown encouraging pathological complete response rates.[11] However, existing evidence primarily derives from controlled populations in prospective clinical trials, and the generalizability of these findings to heterogeneous patient populations in real-world clinical practice—encompassing varying performance statuses, comorbidities, and treatment adherence—remains unclear. Currently, systematic retrospective studies specifically investigating the real-world efficacy and safety of sintilimab in the neoadjuvant treatment of Chinese patients with triple-negative breast cancer are still relatively scarce.

Therefore, this study aims to evaluate the efficacy and safety of neoadjuvant chemotherapy combined with the domestic PD-1 inhibitor sintilimab in the treatment of triple-negative breast cancer within real-world clinical practice, through a single-center, small-sample retrospective cohort study. We hope to provide important real-world evidence-based medical data to support the application of sintilimab in the neoadjuvant setting for triple-negative breast cancer, thereby offering a reference for clinical treatment decisions and the design of subsequent prospective studies.

## Materials and Methods

### 1. Data collection

Employing a retrospective cohort study design and strictly adhering to the predefined inclusion and exclusion criteria, breast cancer patients who underwent neoadjuvant therapy at The First Hospital of Lanzhou University between July 2024 and July 2025 were selected. Their general information, clinicopathological data, and hematological indices were collected. Ultimately, a total of 61 patients with TNBC were included in this study. Based on the use of PD-1 inhibitors during neoadjuvant therapy, the patients were divided into two groups: the PD-1 inhibitor combined with neoadjuvant chemotherapy group (n=27) and the neoadjuvant chemotherapy-alone group (n=34). This study was approved by the hospital’s ethics committee (Approval No. LDYYLL2026-37).

Inclusion criteria: (1) Patients with pathologically confirmed triple-negative breast cancer (TNBC); (2) Breast cancer patients with stage IIA to IIIC disease not amenable to direct surgery; (3) Normal results in blood tests, liver and kidney function tests, and electrocardiogram (ECG), meeting the criteria for antitumor therapy; (4) No prior local treatment before neoadjuvant therapy; (5) Non-pregnant and non-lactating women; (6) Patients with an Eastern Cooperative Oncology Group (ECOG) performance status of 0 to 1. Exclusion criteria: (1) Patients with other concurrent primary malignancies; (2) Patients with incomplete clinical records; (3) Patients with any contraindications to immunotherapy.

### 2. Neoadjuvant Treatment Regimens and Drug Administration

PD-1 Inhibitor: Sintilimab 200 mg, administered via intravenous infusion on day 1 of a 21-day cycle, for 6 to 8 cycles.

Chemotherapy Regimens:

Modified KEYNOTE-522 Regimen: A regimen consisting of 4 cycles of nab-paclitaxel combined with carboplatin, followed by 4 cycles of an anthracycline (such as doxorubicin hydrochloride liposome, pirarubicin hydrochloride, or epirubicin hydrochloride) combined with cyclophosphamide. Nab-paclitaxel + Carboplatin:

Nab-paclitaxel 220 mg/m² combined with carboplatin dosed at an area under the curve (AUC) = 5, administered via intravenous infusion on day 2 of a 21-day cycle, for 4 cycles. Anthracycline + Cyclophosphamide: Doxorubicin hydrochloride liposome (CSPC) 30 mg/m², or pirarubicin hydrochloride (Joint Venture) 45 mg/m², or epirubicin hydrochloride for injection (Joint Venture) 100 mg/m², combined with cyclophosphamide (Imported) 500 mg/m², administered via intravenous infusion on day 2 of a 21-day cycle, for 4 cycles.

TCb Regimen: A regimen consisting of 6 cycles of nab-paclitaxel combined with carboplatin. Nab-paclitaxel + Carboplatin: Nab-paclitaxel 175 mg/m² combined with carboplatin dosed at AUC = 5, administered via intravenous infusion on day 2 of a 21-day cycle, for 6 cycles.

During treatment, any discomfort was to be promptly reported to the supervising physician for immediate symptomatic management. For all patients, systemic laboratory tests and imaging examinations were performed starting 3 weeks after completion of neoadjuvant therapy, followed by breast-conserving surgery or modified radical mastectomy based on the clinical assessment.

### 3. Detection of Relevant Biomarkers

Prior to neoadjuvant chemotherapy (NAC), the expression of estrogen receptor (ER), progesterone receptor (PR), human epidermal growth factor receptor 2 (HER-2), and Ki-67 in breast target tissues was assessed by immunohistochemistry (IHC). Both ER and PR were considered negative if their expression levels were <1%. HER-2 positivity was defined as an IHC result of (+++) or (++) confirmed by a positive fluorescence in situ hybridization (FISH) test; all other cases were classified as HER-2 negative. Fasting peripheral venous blood samples were collected from patients in the morning on two occasions: the day before the first chemotherapy cycle and on the third day after the completion of the final chemotherapy cycle.Complete blood counts (including white blood cell count [WBC], hemoglobin concentration [Hb], and platelet count [Plt]), liver function indices (including aspartate aminotransferase [AST] and alanine aminotransferase [ALT]), and tumor markers (including carcinoembryonic antigen [CEA], cancer antigen 125 [CA-125], and cancer antigen 153 [CA-153]) were measured for all patients before and after treatment in the clinical laboratory of our hospital. All detection procedures and instrumentation adhered to standard operating protocols and underwent regular quality control.

### 4. Efficacy Evaluation Criteria

#### 4.1 Clinical Efficacy Evaluation

Clinical efficacy was assessed according to the Response Evaluation Criteria in Solid Tumors (RECIST) version 1.1. Definitions are as follows:Complete Response (CR): Disappearance of all target and non-target lesions.Partial Response (PR): At least a 30% decrease in the sum of diameters of target lesions, with no progression of non-target lesions.Progressive Disease (PD): At least a 20% increase in the sum of diameters of target lesions, or the appearance of one or more new lesions.Stable Disease (SD): Neither sufficient shrinkage to qualify for PR nor sufficient increase to qualify for PD.Based on these categories, the following rates were calculated:Objective Response Rate (ORR) = (Number of patients with CR + PR) / Total number of patients ×100%.Disease Control Rate (DCR) = (Number of patients with CR + PR + SD) / Total number of patients × 100%.

#### 4.2 Postoperative Pathological Evaluation

Postoperative pathological assessment was conducted using the Miller–Payne Grading (MPG) system and the Residual Cancer Burden (RCB) system. Based on this assessment, responses were categorized as either pathological complete response (pCR) or non-pCR. According to the *Expert Consensus on Pathological Diagnosis after Neoadjuvant Chemotherapy for Breast Cancer*, pCR was defined as the absence of invasive carcinoma in the primary breast site (the presence of ductal carcinoma *in situ* was allowed) combined with negative regional lymph nodes. Specifically, this corresponded to an MPG score of 5 for the primary tumor with no lymph node metastasis, or an RCB score of 0.In the evaluation of efficacy, the pCR rate served as the primary endpoint, while the ORR was a secondary endpoint.

### 5. Adverse Reaction Observation and Evaluation

Adverse reactions observed in patients were evaluated according to the Common Terminology Criteria for Adverse Events (CTCAE) version 5.0. The evaluation period spanned from the first administration of the study treatment to one month after the last dose, with assessments conducted monthly. Adverse reactions were graded for severity from 1 to 5:Grade 1 (Mild):The adverse reaction is largely asymptomatic or causes minimal symptoms, requiring no intervention.Grade 2 (Moderate):The adverse reaction may limit age-appropriate instrumental activities of daily living, and minimal, local, or non-invasive intervention is indicated.Grade 3 (Severe):The adverse reaction is medically significant but not immediately life-threatening, potentially causing significant disability or incapacity, and typically requires hospitalization or prolongation of hospitalization.Grade 4 (Life-threatening):The adverse reaction is life-threatening and necessitates urgent medical intervention.Grade 5 (Death):The adverse reaction directly results in death.

### 6. Statistical Analysis

Data were analyzed using SPSS version 31. A significance level of P<0.05 was defined as statistically significant. Continuous variables are presented as mean ± standard deviation (SD) or median (interquartile range), with comparisons between groups performed using the Student’s t-test or the Mann-Whitney U test, as appropriate. Categorical variables are described as frequency (percentage), and differences between groups were assessed using the chi-square test or Fisher’s exact test.

## Result

### 1. Baseline Clinical Characteristics of Patients

Strict adherence to the inclusion and exclusion criteria resulted in the enrollment of 61 patients with TNBC in this study. The baseline characteristics of the 27 patients who received PD-1 inhibitor combined with neoadjuvant chemotherapy were compared with those of the 34 patients who received neoadjuvant chemotherapy alone. Statistical analysis revealed a statistically significant difference between the two groups in terms of age and chemotherapy regimen (P < 0.05). No significant differences were observed for the other baseline characteristics (P > 0.05). The detailed data are presented in Table 1.

**Table 1.**
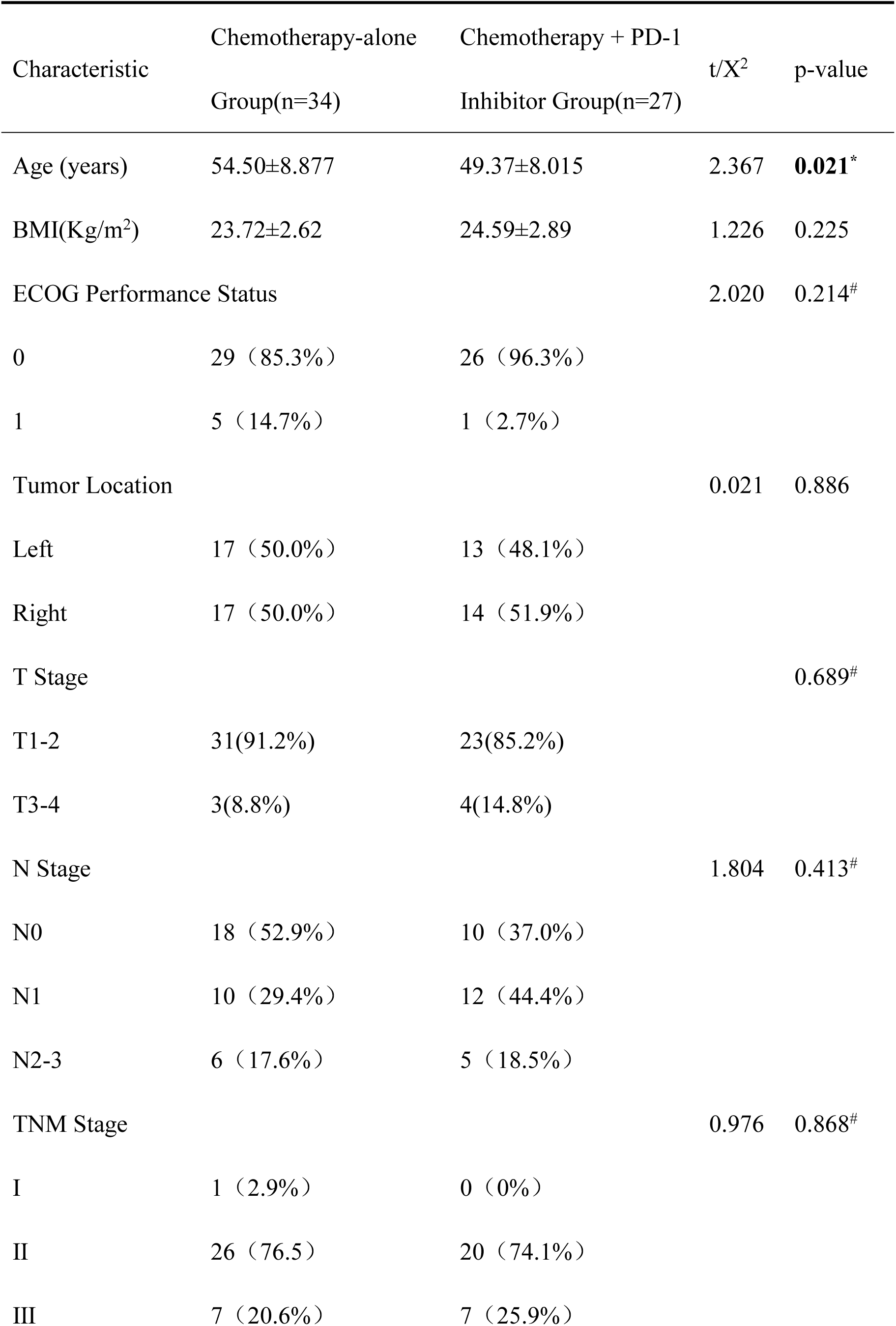
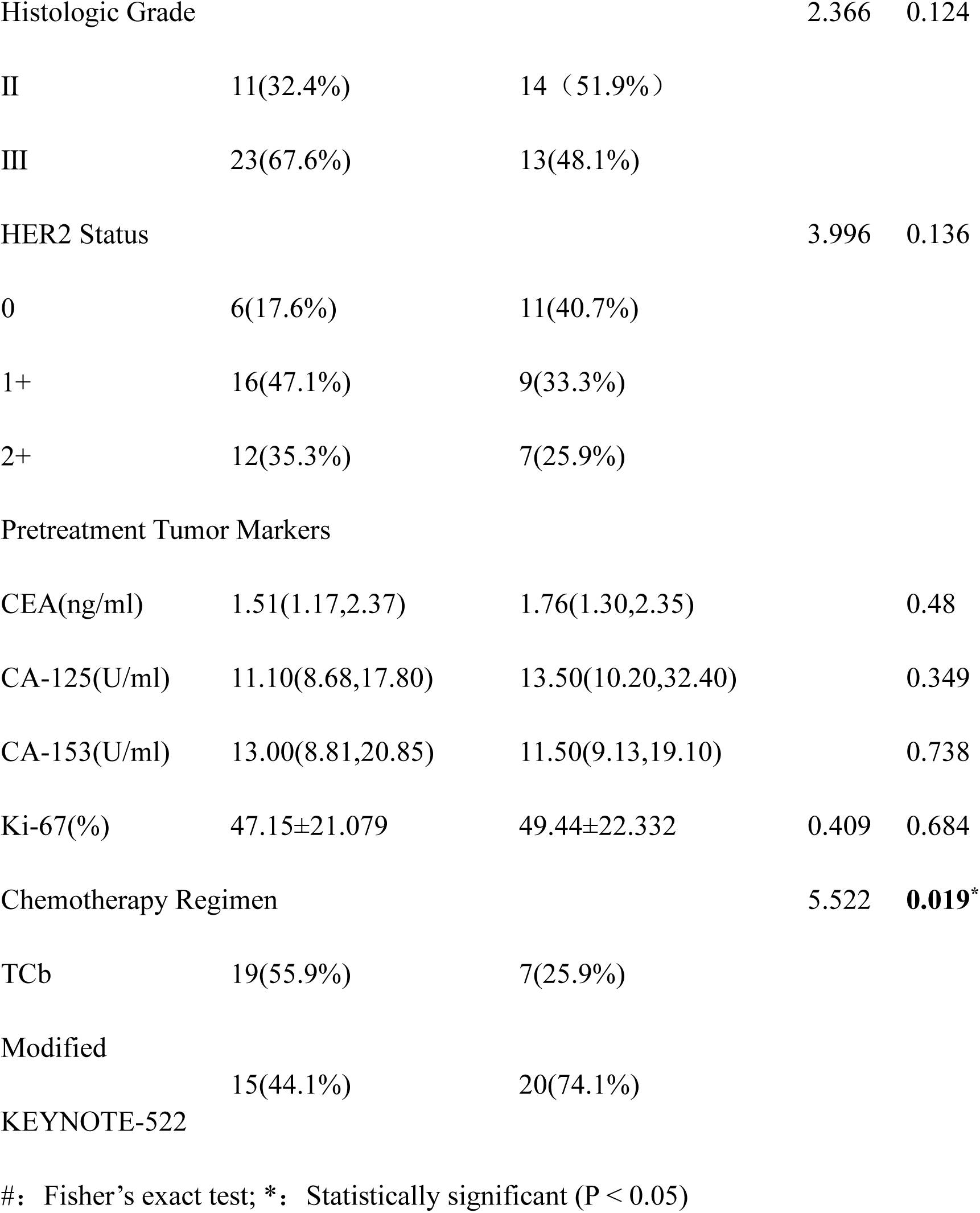
Comparison of Baseline Characteristics Between the Two Groups.

### 2. Efficacy Evaluation

The PD-1 inhibitor combined with neoadjuvant chemotherapy group demonstrated significantly higher rates of pathological complete response (pCR) and objective response rate (ORR) compared to the neoadjuvant chemotherapy-alone group, with the differences being statistically significant (P < 0.05). Detailed data are presented in Table 2.

**Table 2.** Comparison of Efficacy Between the Two Groups.

| Characteristic | Chemotherapy-alone<br>Group(n=34) | Chemotherapy +<br>PD-1 Inhibitor<br>Group(n=27) | t/X<sup>2</sup> | p-value |
| --- | --- | --- | --- | --- |
| Clinical Efficacy Evaluation |  |  | 5.152 | 0.147<sup>#</sup> |
| CR | 1(2.9%) | 2 (7.4%) |  |  |
| PR | 19(55.9%) | 21(77.8%) |  |  |
| SD | 8(23.5%) | 2 (7.4%) |  |  |
| PD | 6(17.7%) | 2(7.4%) |  |  |
| Objective Response<br>Rate | 20(58.8%) | 23(85.2%) | 5.028 | **0.025\*** |
| Disease Control<br>Rate | 28(82.4%) | 25(92.6%) |  | 0.283<sup>#</sup> |
| pCR rate | 11(32.4%) | 16(59.3%) | 4.416 | **0.036\*** |
| Breast-conserving<br>surgery rate | 5(14.7%) | 8(29.6%) | 1.999 | 0.157 |

#: Fisher's exact test; \*: Statistically significant (P < 0.05)

### 3. Comparison of Tumor Markers Between the Two Groups Before and After Treatment

Before treatment, no significant difference was observed in tumor markers between the two patient groups (P > 0.05), with detailed data provided in Table 1. After treatment, the PD-1 inhibitor combined with neoadjuvant chemotherapy group demonstrated significantly lower Ki-67 levels compared to the neoadjuvant chemotherapy-alone group, and the difference was statistically significant (P < 0.05). Detailed data are presented in Table 3.

**Table 3.** Comparison of Post-Treatment Tumor Markers, Hematological Indices, and Liver Function Indices Between the Two Groups.

| Characteristic | Chemotherapy-alone<br>Group(n=34) | Chemotherapy + PD-1<br>Inhibitor Group(n=27) | t/X<sup>2</sup> | p-value |
| --- | --- | --- | --- | --- |
| Post-treatment Tumor Markers |  |  |  |  |
| CEA(ng/ml) | 1.73(1.36,3.16) | 1.83(1.15,3.05) |  | 0.223 |
| CA-125(U/ml) | 8.93(7.86,12.95) | 11.00(10.06,15.75) |  | 0.065 |
| CA-153(U/ml) | 19.12±7.39 | 16.88±7.95 | 1.234 | 0.222 |
| Ki-67(%) | 35.00(5.00,55.00) | 5.00(5.00,30.00) |  | **0.042\*** |

#: Fisher's exact test; \*: Statistically significant (P < 0.05)

### 4. Comparison of Adverse Reactions Between the Two Groups

Adverse reactions were assessed using CTCAE version 5.0. No statistically significant difference was observed in the overall incidence of adverse reactions between the two groups (P > 0.05). A further comparison of the incidence rates for individual adverse events revealed that, aside from leukopenia which occurred at a significantly higher rate in the PD-1 inhibitor combination group compared to the chemotherapy-alone group (P < 0.05), there were no other statistically significant differences (P > 0.05), as detailed in Table 4. During the treatment period, most adverse reactions were grade 1 or 2 in severity. Events such as hepatic impairment, nausea/vomiting, fatigue, leukopenia, and peripheral neuropathy were relatively frequent. All patients showed marked improvement following symptomatic management, with no patient discontinuing treatment prematurely.

**Table 4.** Comparison of Adverse Reactions Between the Two Groups.

| Characteristic | Chemotherapy-alone<br>Group(n=34) | Chemotherapy +<br>PD-1 Inhibitor<br>Group(n=27) | t/X<sup>2</sup> | p-value |
| --- | --- | --- | --- | --- |
| Adverse Reactions | 30(88.2%) | 26(96.3%) |  | 0.371<sup>#</sup> |
| Hepatic<br>Impairment | 10(29.4%) | 9(33.3%) | 0.108 | 0.743 |
| Fever | 1(2.9%) | 0(0.0%) |  | 1.00<sup>#</sup> |
| Anemia | 2(5.9%) | 4(14.8%) |  | 0.392<sup>#</sup> |
| Leukopenia | 7(20.6%) | 14(51.9%) | 6.516 | **0.011<sup>\*</sup>** |
| Thrombocytopenia | 4(11.8%) | 5(18.5%) |  | 0.492<sup>#</sup> |
| Diarrhea | 0(0.0%) | 0(0.0%) | - | - |
| Fatigue | 19(55.9%) | 9(33.3%) | 3.081 | 0.079 |
| Hypothyroidism | 1(2.9%) | 0(0.0%) |  | 1.00<sup>#</sup> |
| Peripheral<br>neuropathy | 21(61.8%) | 40.7(%) | 2.667 | 0.102 |

#: Fisher's exact test; \*: Statistically significant (P < 0.05)

## Discussion

This retrospective cohort study revealed that among patients with triple-negative breast cancer receiving neoadjuvant therapy, the addition of the PD-1 inhibitor sintilimab to chemotherapy significantly improved the rates of pathological complete response (pCR) and objective response (ORR) compared to chemotherapy alone. Furthermore, it effectively reduced the levels of the tumor cell proliferation marker Ki-67, with an overall manageable safety profile. These findings contribute real-world clinical practice evidence supporting the application of sintilimab in the neoadjuvant treatment of TNBC.

In this study, the pCR rate of 59.3% achieved in the combination therapy group is particularly noteworthy. This value falls within a range comparable to the pCR rate (64.8%) reported for pembrolizumab combined with chemotherapy in the KEYNOTE-522 trial, [5] suggesting that different PD-1 inhibitors may harbor similar potential for enhancing the efficacy of neoadjuvant therapy in TNBC.Pathological complete response (pCR) serves as a robust surrogate endpoint for event-free survival and overall survival following neoadjuvant therapy in triple-negative breast cancer (TNBC). [12] Therefore, the significant increase in the pCR rate observed in this study (59.3% vs. 32.4%) holds important clinical prognostic value and may translate into long-term survival benefits for patients. Furthermore, the significantly higher ORR in the combination therapy group (85.2% vs. 58.8%) further confirms the advantage of this regimen in inducing early tumor regression. This aligns with the mechanism of immunotherapy, which activates the host immune system to generate sustained anti-tumor effects.

The significant reduction in the Ki-67 index provides biological rationale supporting the antitumor activity of the combination regimen. Ki-67 is a well-established marker of cellular proliferation, whose expression levels are associated with tumor aggressiveness and poor prognosis. The extent of Ki-67 decline after neoadjuvant therapy has been correlated with improved treatment response and survival outcomes. [13] In this study, the greater reduction in Ki-67 observed in the combination therapy group not only correlates with the higher pCR rate, but also indirectly reflects that the combination of immunotherapy and chemotherapy may more effectively inhibit tumor proliferative activity and alter the biological behavior of the tumor.

Regarding safety, the combination therapy group exhibited a manageable toxicity profile. Although the incidence of leukopenia was increased—likely related to the combined effects of known chemotherapy-induced myelosuppression and potential synergistic impacts of immunotherapy—the incidence rates of other common adverse events, such as hepatic dysfunction, fatigue, and hypothyroidism, were not significantly elevated. This finding is largely consistent with the established safety characteristics of most PD-1/PD-L1 inhibitors combined with chemotherapy, which primarily increase the risk of immune-related adverse events and specific hematologic toxicities without substantially altering the overall profile of chemotherapy’s inherent toxicities. [14] This suggests that, with adequate monitoring and management, the application of this combination regimen in real-world clinical practice is feasible.

However, this study has several limitations. Firstly, its retrospective design cannot completely avoid selection bias and potential confounding factors. Secondly, the overall sample size is limited (N=61), with a particularly small cohort in the combination therapy group (n=27). This may be insufficient to adequately capture rare adverse events and restricts the statistical power for conducting in-depth subgroup analyses and multivariable testing. Thirdly, constrained by the retrospective data, this study lacked systematic assessment of PD-L1 expression status and other key tumor immune microenvironment features (such as CD8⁺ T-cell density and tumor-infiltrating lymphocyte levels). Consequently, it was unable to explore potential correlations between these biomarkers and clinical efficacy. Finally, the primary endpoint employed (e.g., pCR rate) is only a surrogate endpoint, and its correlation with long-term clinical benefits (such as event-free survival and overall survival) remains to be confirmed.

Furthermore, the assessment of delayed immune-related adverse events might be inadequate.Future research should focus on conducting large-scale, prospective, multicenter clinical trials to validate the current findings. It should also delve deeper into exploring predictive biomarkers, including tumor mutational burden and immune cell subset characteristics, with the aim of enabling biomarker-guided precision treatment strategies.

## Conclusion

In conclusion, the real-world data from this study indicate that adding sintilimab to a standard neoadjuvant chemotherapy regimen significantly improved the pathological complete response rate and objective response rate in patients with locally advanced triple-negative breast cancer. It also effectively suppressed tumor proliferative activity without a significant increase in the overall toxicity risk. These findings provide preliminary clinical evidence supporting sintilimab as a potential strategy in the neoadjuvant treatment of TNBC. This study also represents an important addition to the real-world efficacy and safety data for the domestic PD-1 inhibitor sintilimab in this field.

## Ethical Statement

The study protocol was approved by the Ethics Committee of The First Hospital of Lanzhou University (approval number LDYYLL2026-37). The informed consent was waived due to its retrospective and non-interventional nature.

## Author Contributions

Conceived and designed the analysis: Gao ZB, Shan BF, ChenXH, Tang JM.

Collected the data: Gao ZB, Liang HW, Bai X, Dong K, Li J, Qiao WJ. Contributed data or analysis tools: Gao ZB.

Performed the analysis: Gao ZB, Dong K. Wrote the paper: Gao ZB.

Supervision: Tang JM, Shan BF, ChenXH.

## Conflicts of Interest

Conflict of interest relevant to this article was not reported.

## Funding

This research did not receive any specific grant from funding agencies in the public, commercial, or not-for-profit sectors.

## Data Availability

All data produced in the present work are contained in the manuscript

## Notes

### Competing Interest Statement

The authors have declared no competing interest.

### Funding Statement

This study did not receive any funding

